# A quantitative approach to evidence triangulation: development of a framework to address rigour and relevance

**DOI:** 10.1101/2024.09.20.24314046

**Authors:** Chin Yang Shapland, Joshua A. Bell, Maria-Carolina Borges, Ana Goncalves Soares, George Davey Smith, Tom R. Gaunt, Deborah A. Lawlor, Luke A. McGuinness, Kate Tilling, Julian P.T. Higgins

## Abstract

Triangulation is an approach to strengthen causal inference by integrating evidence from multiple sources. Most studies using triangulation have qualitatively examined whether different studies agree upon the presence of a causal effect, rather than estimated the effect by quantitatively integrating results. Here, we develop a framework for quantitative triangulation. We first address how to relate study specific research questions to an overall target causal question (relevance) and then assess the directions and magnitudes of bias in each study (rigour), before combining the results using meta-analysis adjusted for the biases.

We illustrate our framework by triangulating evidence from randomized controlled trials (RCTs), Mendelian randomization (MR) and conventional multivariable regression (MVR) to estimate the effect of beta-carotene on coronary heart disease (CHD) and cardiovascular disease (CVD). Five RCTs and one MR study showed little evidence of a causal relationship between beta-carotene and CHD (relative risk (RR)=1.00 with 95% CI=0.98 to 1.01 and RR=1.02 with 95% CI=0.98 to 1.07, respectively). Thirteen MVR studies indicated that high intake of beta-carotene reduces CHD risk (RR=0.83 with 95% CI=0.76 to 0.91). After applying our framework, the three study designs agreed that there is no strong evidence against the null hypothesis that beta-carotene intake does not affect risk of CHD (RR=0.99 with 95% CI=0.82 to 1.21). Findings were similar for CVD.

Our framework shows how to address rigour and relevance quantitatively when triangulating evidence from different study designs. We highlight the importance of explicitly defining the target and study-specific research questions.

## Introduction

Triangulation of epidemiological evidence has been defined as “*the practice of strengthening causal inferences by integrating results from several approaches, where each approach has different (and assumed to be largely unrelated) key sources of potential bias*” [1]. The motivation is that where studies are subject to different types of biases, if the conclusions from all studies suggest that there is a causal effect of exposure on outcome, we have added confidence that there is a true causal effect. Increasing interest in triangulation is accompanying the widening of available approaches for evaluating effects of exposures, such as genetics-based methods (e.g. Mendelian randomization (MR)) [2] and target trial emulation analyses [3].

Applications of triangulation have mostly taken a ‘qualitative’ approach, seeking primarily to establish the presence and direction of a causal relationship. Here, we develop a quantitative approach, aiming to estimate the magnitude of the causal relationship. Quantifying relationships is important for several reasons, including identification of safe levels of an exposure, examining trade-off of benefits and harms, and evaluation of interventions.

To quantify a relationship, the parameter(s) being estimated must be defined. This requires specification of the research question (the ‘target question’) in terms of the population, exposure and outcome of interest and the assumed nature of the relationship between the exposure and the outcome (e.g. linear or non-linear). Individual studies assembled for a triangulation exercise will rarely each provide an answer to the target question directly, for two reasons. First, the participants, exposures and outcomes will typically differ from each other and from the target question – referred to as ‘relevance’ or ‘external bias’ [4]. Second, the studies will suffer from different biases – referred to as ‘rigour’ or ‘internal bias’ [4].

The aim of this paper is to develop and illustrate a framework for quantitatively triangulating evidence from studies taking different approaches to answer a target question. To put the triangulation problem into a quantitative framework, we additionally assume that the inherent biases of the included studies are not all in the same direction. Under this assumption, convergence of the bias-adjusted estimates increases confidence that the resulting estimate is closer to the target causal effect [5]. We use the effect of beta-carotene on cardiovascular disease (CVD) risk as a case study, as previous research highlighted differences between randomized controlled trials (RCTs) and observational estimates [6]. To deal with differences in relevance (external bias), we transform or scale the results of the studies to match the target question, based on a combination of assumptions and external data.

We deal with rigour (internal bias) by quantifying the likely biases, based on formal assessments of risk of bias using standard tools [7–10]. We undertake the analysis within a Bayesian framework, using the quantified bias assessments as informative prior distributions to perform bias adjustments [4]. Intuitively, the aim of the quantitative triangulation analysis is to reduce the between-study heterogeneity by adjusting for study-specific biases. We therefore do not exclude studies judged at “high” or “very high” risk of bias, although the prior distributions for the bias for these studies have large variance so they (appropriately) have a high degree of uncertainty. Nonetheless, if a study is deemed to be completely flawed (e.g. used a method that has subsequently been retracted), it should be excluded.

## Methods

We develop our framework in terms of: defining the target causal question; identifying evidence; assessing relevance of individual studies to this target question; assessing biases in the individual studies; judging heterogeneity of bias between approaches and between studies; and finally combining evidence from all studies whilst adjusting for bias and relevance (as shown in Figure 1).

**Figure 1:**
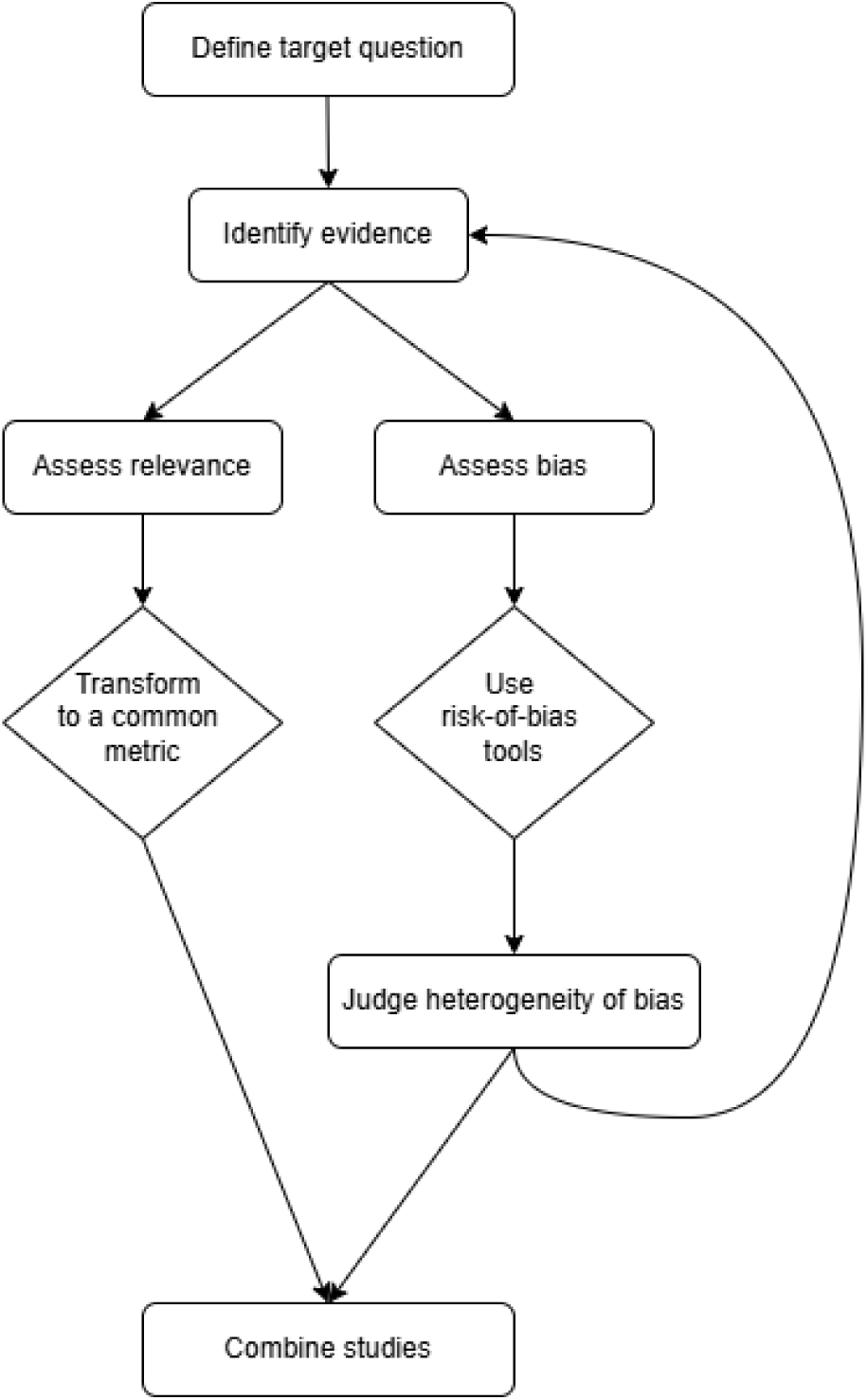
Flow chart demonstrating our proposed triangulation framework.

### Defining the target causal question

Defining the target causal question is the important first step of triangulation (as in the target trial emulation framework [3]). This step enables us to assess the relevance of each study’s causal question and map the results to a common metric. We consider six key aspects in defining the target question: the population, exposure measure, exposure window, how exposure is to be summarized over time (e.g. single exposure event, average exposure over mid-life, etc), outcome measure, and the statistical estimand [11]. Ideally, as with any study design, the causal question should be specified before identifying studies to be included. eTable 1 lists some considerations for each of these aspects of the target causal question.

### Identifying evidence

Our triangulation framework assumes that the included studies to answering the target question have biases in different directions. Note that this may only become confirmed once the risk-of-bias assessments have been performed, as discussed below. One way to achieve different directions of bias might be to use different study designs. For example, biases in RCTs (e.g. loss to follow-up bias) will differ from biases in observational studies (e.g. unmeasured confounding). Use of different study designs is not a prerequisite, however. Triangulation can still proceed within a study design if studies are likely to have varying directions of bias; for example, across observational studies undertaken in different settings where the direction of unmeasured confounding might differ [5].

### Relevance (external bias): assessing and converting results to a comparable metric

The next step is to assess the ‘relevance’ of the included studies, i.e. how directly each individual study answers the target causal question. First, we need to determine the research question addressed by each study, in terms of the same six aspects that define the target question. The population and outcome are given by the eligibility criteria and outcome measurements used in the study. The exposure measure and duration in RCTs are straightforward to extract from the study description. Defining the exposure of interest for observational studies is more challenging because the assumed start time and exposure window are rarely articulated and may need to be deduced. For example, a study relating body mass index (BMI) at baseline in UK Biobank to later risk of CVD [12] may implicitly be considering average BMI during middle age as the exposure.

Once the research question for each study has been identified, final decisions about inclusion of each study can be made. Subjective decisions include whether to incorporate studies using different exposures (e.g. percentage of fat mass instead of BMI), outcomes (e.g. blood pressure instead of CVD) or in different populations. One approach for defining the research question would be to use the target trial emulation framework, which suggests defining the intervention for an “ideal” RCT [3] and relevance of different potential interventions to this target intervention is then a subjective decision (e.g. the equivalence of the effect on CVD of BMI changes via drugs, dietary or exercise interventions).

To be included in a quantitative triangulation, the effect estimated by a study must be mathematically relatable to the effect defined by the target question. Such a mathematical relationship could use information from an external source, providing the relationship is considered transportable to the study under consideration. For example, consider a target question is the effect of BMI on subsequent depression. Studies relating the percentage of fat mass to subsequent depression could be included by using external information about the relationship between BMI and percentage of fat mass, and assuming that an intervention on fat mass would produce the same effect on depression as an intervention on BMI. A study relating self-reported body shape (underweight, normal, overweight) to depression might not be included even if there was external information to relate BMI to these categories, because it is likely that self-reported body weight has pathways to depression other than through measured BMI (e.g. an intervention that changed self-reported body shape might have a different effect on depression from an intervention to change BMI).

### Rigour (internal bias): risk-of-bias assessment

Different study designs rely on different casual assumptions [5], summarized in Table 1. Our framework uses rigorously developed tools, rooted in these causal assumptions, to assess risk of bias in each study. Examples of such tools are RoB 2 [7], ROBINS-I [8], ROBINS-E [9] and the tool developed by Mamluk *et al.* [10] to assess the risk of bias in RCTs, conventional observational studies of interventions and exposures and MR studies, respectively. Different tools have different bias domains and levels of risk-of-bias judgements. Table 2 illustrates the bias domains assessed and risk-of-bias judgement options for three of these tools. Note that the domains are labelled for convenience, but there is no correspondence in domains across study types. Standard systematic review good practice should be followed, including having at least two independent assessors of risk of bias for each study.

**Table 1:**
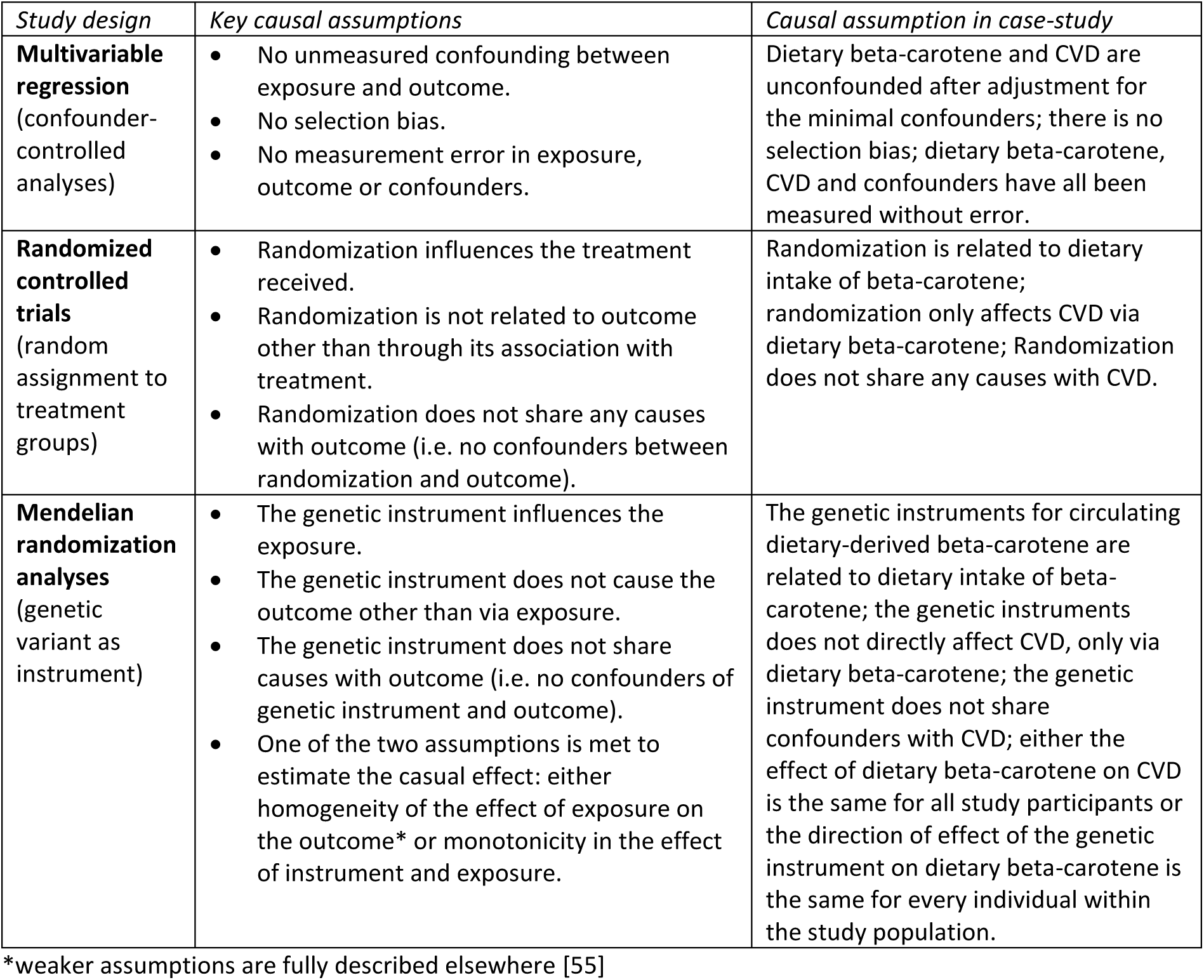
Summary of key assumptions for causal inference in used study design.

| *Study design* | *Key causal assumptions* | *Causal assumption in case-study* |
| --- | --- | --- |
| **Multivariable regression**<br>(confounder-controlled analyses) | No unmeasured confounding between exposure and outcome.<br>No selection bias.<br>No measurement error in exposure, outcome or confounders. | Dietary beta-carotene and CVD are unconfounded after adjustment for the minimal confounders; there is no selection bias; dietary beta-carotene, CVD and confounders have all been measured without error. |
| **Randomized controlled trials**<br>(random assignment to treatment groups) | Randomization influences the treatment received.<br>Randomization is not related to outcome other than through its association with treatment.<br>Randomization does not share any causes with outcome (i.e. no confounders between randomization and outcome). | Randomization is related to dietary intake of beta-carotene; randomization only affects CVD via dietary beta-carotene; Randomization does not share any causes with CVD. |
| **Mendelian randomization analyses**<br>(genetic variant as instrument) | The genetic instrument influences the exposure.<br>The genetic instrument does not cause the outcome other than via exposure.<br>The genetic instrument does not share causes with outcome (i.e. no confounders of genetic instrument and outcome).<br>One of the two assumptions is met to estimate the casual effect: either homogeneity of the effect of exposure on the outcome* or monotonicity in the effect of instrument and exposure. | The genetic instruments for circulating dietary-derived beta-carotene are related to dietary intake of beta-carotene; the genetic instruments does not directly affect CVD, only via dietary beta-carotene; the genetic instrument does not share confounders with CVD; either the effect of dietary beta-carotene on CVD is the same for all study participants or the direction of effect of the genetic instrument on dietary beta-carotene is the same for every individual within the study population. |

\*weaker assumptions are fully described elsewhere [55]

**Table 2:** Bias domains and risk-of-bias judgements for each study design.

| *Study Design* | *Randomized controlled trials* | *Conventional observational studies of exposure effects* | *Mendelian randomization* |
| --- | --- | --- | --- |
| *Tools* | *RoB 2* | *ROBINS-E* | *Mamluk et al. [10]* |
| *Bias domains* |  |  |  |
| D1 | Randomization process | Confounding | Weak instrument |
| D2 | Deviations from intended interventions | Measurement of the exposure | Genetic confounding |
| D3 | Missing outcome data | Selection of participants into the study (or into the analysis) | Other confounding |
| D4 | Measurement of the outcome | Post-exposure interventions | Additional direct effects between instrumental variable and outcome (exclusion restriction assumption) |
| D5 | Selection of the reported result | Missing data | Selection of participants |
| D6 | - | Measurement of outcomes | - |
| D7 | - | Selection of the reported result | - |
| *Risk-of-bias judgement options* |  |  |  |
|  | Low / Some concerns / High | Low / Some concerns / High / Very high | Low / Moderate / High |

RoB 2: Risk of Bias 2, ROBINS-E: Risk Of Bias In Non-Randomized Studies tools for observational studies of exposure effects.

### Judging heterogeneity of bias

For triangulation, the studies included should be vulnerable to different biases, resulting in different directions of bias. In our quantitative framework, we assess this by predicting the magnitude and direction of bias in each study. Evidence to inform this is gathered through the risk-of-bias assessments. Variation in biases would manifest itself in different profiles of risk of bias across the bias domains and different predicted directions of bias for at least some of the main domains.

The first step in quantifying the bias is to postulate the type of bias in each domain, and whether the bias is likely to be additive or proportional. An additive bias either “Favours experimental” or “Favours comparator”. Proportional bias acts multiplicatively on the estimated exposure effect so is either “Towards the null” or “Away from the null”. Bias is classified as “Unpredictable” if the direction of bias cannot be predicted. To exemplify these considerations, consider plausible bias in the effect of statin use (versus no medication) on Alzheimer’s disease. If older people are more likely to be prescribed statins, and a study does not adjust for age this will likely induce an additive bias in favour of no medication. If a study does not provide an analysis protocol, the authors might have chosen results that favour a particular direction of effect, thereby introducing additive bias towards the desired direction. Alternatively, authors may favour the most “statistically significant” results, thereby introducing a multiplicative bias (i.e. the result selected for publication is that furthest from the null in either direction). For formal definitions of types and directions of bias see Turner *et al.* [4]. Algebraic details of the additive and proportional biases are given in eAppendix 1b. Note that a given bias may act differently in different studies or contexts. Careful consideration of the process of each bias is needed to decide on the form and order to apply bias adjustments [13].

The final step to quantify the likely extent of bias (based on judgements in Table 2), is to specify prior distributions for bias, which capture its type (additive or proportional) and direction for each study. For “unpredictable” bias, the prior distribution may have a mean of zero with a non-zero variance – thus “unpredictable” bias does not change the effect estimate but increases its uncertainty. We do not provide general guidance on specification of priors, as this will differ for each case, although we present some considerations in the discussion.

At this stage, the supposition that biases vary in direction can be confirmed. If there is no variation in bias, the researcher may need to revisit the “Identifying evidence” step before proceeding with a triangulation, by finding further sources of evidence (for example in another population or using a different study design) or possibly carry out a new study.

### Combining relevant studies

We use the *triangulate* R package to combine bias-adjusted effect estimates for each study [14]. The *triangulate package* provides a systematic way to integrate assessments from risk-of-bias tools and adjusts for bias and relevance concerns from multiple study designs using the prior distributions derived above, then combines the adjusted estimates using standard meta-analytic methods. A key feature of our triangulation framework is the assessment of heterogeneity using statistics such as I^2^, which quantifies the proportion of variability in effect estimates that is attributable to between-study heterogeneity rather than sampling error [15]. We assume that part of the heterogeneity observed between study designs is explained by variation in study design-specific biases. Therefore, if these biases are correctly identified and modelled, I^2^should decrease following bias adjustment. Such a reduction in heterogeneity would be consistent with a reduction in bias-related variation between studies and, consequently, increase confidence in the converged effect estimate. Furthermore, we do not assume that the bias-adjusted estimates are perfectly unbiased; rather, we assume that they are unbiased on average. In contrast, conventional meta-analysis assumes that the observed study estimates are unbiased on average.

## Application to case-study

### Defining the target causal question

To illustrate our framework, we investigate the effect of dietary beta-carotene intake on CVD risk. We defined our target population as UK adults over 40 years of age (since risk of CVD increases exponentially after age 40 [16]) who are initially free from known CVD (since occurrence of a CVD event may change dietary habits). We considered an intervention resulting in an average and sustained increase of 5,000 µg in dietary beta-carotene per day, with follow-up starting at initiation of this intervention [17]. Our main outcome is any CVD, including both fatal and non-fatal events. A secondary outcome is coronary heart disease (CHD), including fatal and non-fatal events [18]. We assumed the risk ratio (RR) for CVD to be constant throughout observed follow-up. Our causal question is therefore: *in the population of UK adults (above age 40) initially free of known CVD, what is the effect of an average increase of 5,000 µg per day of dietary beta-carotene on the incidence of a CVD [or CHD] event over the period of observed follow-up, expressed as a RR assumed constant?*

### Identifying evidence

We searched the Scopus database in February 2023 to identify systematic reviews of the effect of beta-carotene on CVD or CHD using the following search terms within title, abstract or keywords: (“vitamin A” or “retinol” or “Provitamin A” or carotene or carotenes or carotenoid or “β-carotene” or “beta-carotene” or “lycopene”) and (“coronary heart disease” or “heart disease” or CHD or “cardiovascular disease” or CVD) and (“meta-analysis” or “systematic review”). We sought the most recent systematic review covering each different study design: RCTs, multivariable regression (MVR) analyses of observational studies and MR analyses. We did not exclude studies of circulating beta-carotene, as we assume that dietary beta-carotene will act via its impact on circulating beta-carotene.

We identified a systematic review of MVR of observational studies and extracted data from the 19 included studies for the risk of CVD and CHD associated with beta-carotene (dietary and circulating) [19]. From a systematic review of RCTs, we extracted data from eight trials of the effect of supplemental beta-carotene on risk of CVD and CHD [20]. We did not identify any systematic reviews of MR studies. Therefore, we searched for individual MR studies of the potential effects of dietary intake of beta-carotene on CVD/CHD using Scopus, as previously, and a similar search but the term “Mendelian randomization” replacing the systematic review terms (search run February 2023). We identified one MR study from 2021 for the effect of circulating beta-carotene on risk of CHD, which included three estimates from three separate populations [21].

eAppendix 2a gives further details of study exclusion criteria and data transformation in the systematic reviews, as well as details of instrument selection, samples for instrument-exposure and instrument-outcome association in the MR study.

### Relevance (external bias): assessing and converting results to a comparable metric

In this section we assess ‘relevance’ in terms of the target population, exposure measure, exposure window, definition of exposure over time (Table 3), outcome measure and the statistical estimand. eTables 2 and 3 give the country, population, age range, follow-up time, exposure and outcome measure of each study.

**Table 3:**
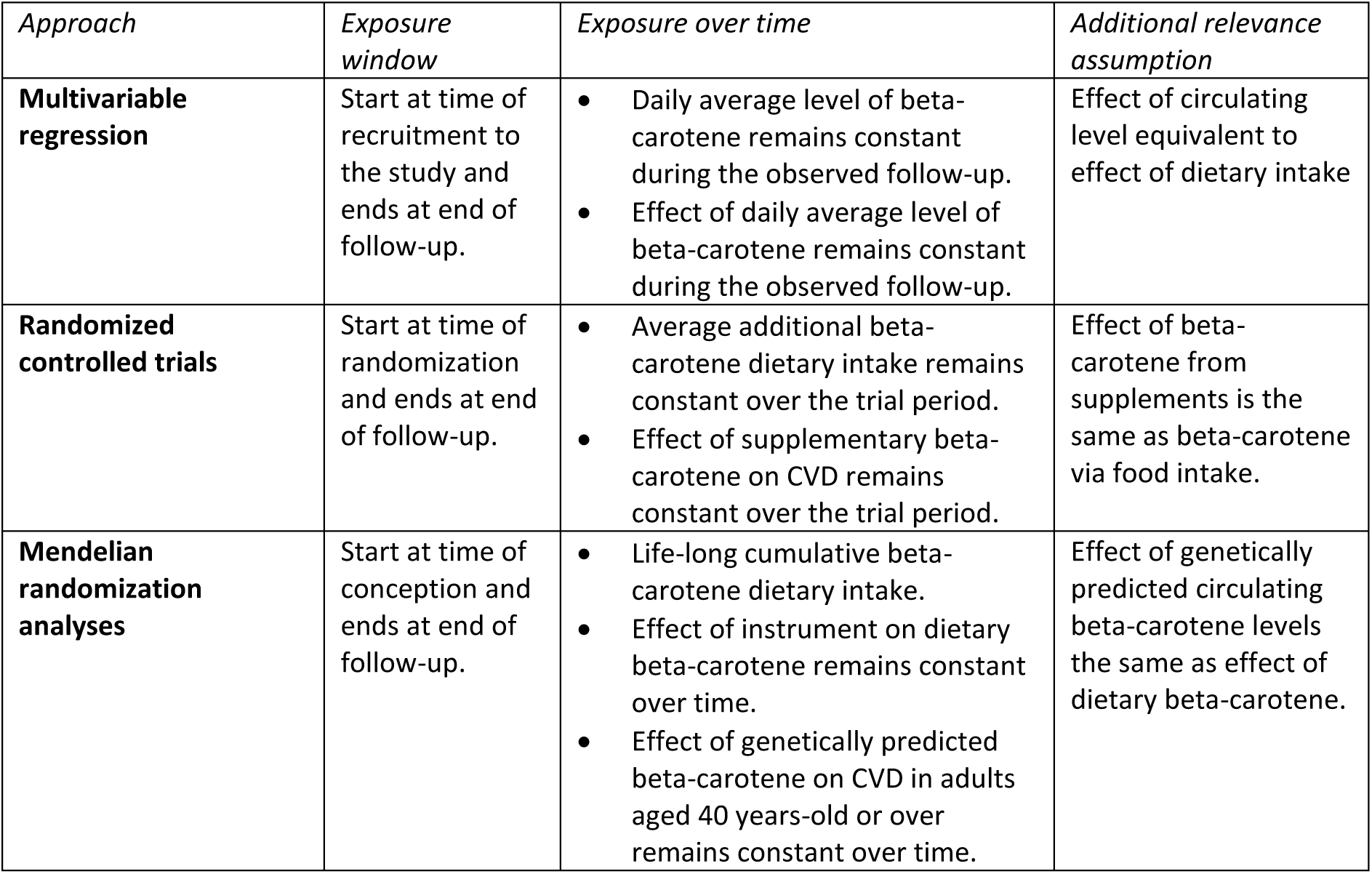
The key assumptions when assessing relevance of exposure from different approaches in case-study.

We first specified the research question for each study. There was no strong prior evidence of heterogeneity in the effect of dietary beta-carotene on risk of CVD/CHD, so we assumed studies were similarly relevant irrespective of their location, or distribution of baseline characteristics such as sex. The exposure measures were either dietary or circulating beta-carotene. However, the assumed exposure window was not articulated in any study. Therefore, we specified the exposure window as starting in each MVR study at the time of recruitment to the study, and starting in each RCT or MR study at the time of randomization, and lasting until the end of follow up. This choice also matched the analyses performed in the studies most closely. eAppendix 2b provides further discussion of the choice of exposure window.

The summary of exposure over time was also not specified in any study. We decided that, for MVR studies, the summary exposure is the daily average beta-carotene dietary intake over time. For RCTs, the summary exposure would be the average additional beta-carotene dietary intake per day during the trial period. For the MR study we considered the summary exposure to be daily beta-carotene dietary intake, as it is assumed that the genetic instrument acts on the exposure from conception/foetal development onwards and has a time-constant effect [22]. Note that the genetic variant could also have time-or age-dependent effects [23], as previously shown for BMI [24].

In each study, the outcome was whether an individual was diagnosed with CVD and CHD defined by International Classification of Diseases either version 9 or 10. Our parameter of interest is the RR for the outcome, given an increase of 5,000 µg per day of dietary beta-carotene sustained over follow-up. As CVD and CHD are rare events, we have assumed that odds ratios, hazard ratios, incidence rate ratios and RRs would be approximately equal.

To map results to a comparable metric, we transformed dosage of exposure level of each study to our pre-specified exposure, “*5,000 µg increase per day of dietary beta-carotene”*. We converted circulating beta-carotene to dietary beta-carotene for the MVR and MR studies that used circulating beta-carotene as their exposure, using results of a meta-analysis of the correlation between dietary and plasma beta-carotene [25]. We assumed this conversion applies to every individual and that dietary beta-carotene intake would only affect outcomes via its effect on circulating beta-carotene. If the conversion is not constant across individuals, the uncertainty of estimates for studies using circulating beta-carotene will be under-estimated. Bias would be introduced if factors affecting the association between dietary and circulating beta-carotene also affect the outcome. For each RCT, we converted the risk ratio for beta-carotene supplements compared to controls to per 5,000 µg/day. See more details in eAppendix 2c.

### Rigour (internal bias): risk-of-bias assessment

We used the tools RoB 2 [7], ROBINS-E[9] and developed by Mamluk *et al.* [10] to assess the risk of bias in RCTs, conventional observational studies and MR studies, respectively. Two assessors independently assessed risk of bias for each study and reached a consensus judgement. Any discrepancies were discussed with a third assessor.

For the assessment of MVR studies using ROBINS-E, after discussions with content experts we agreed on a minimal set of confounders to be included (eTable 5), without which the study would be judged to be at “Very high risk of bias” for Domain 1 (Risk of bias due to confounding). This minimal set of confounders was age, sex, socioeconomic background, ethnicity, BMI and dietary features (including vitamins via food and supplements or dietary intake measured as energy intake).

### Judging heterogeneity of bias

We included three different types of study design, which inherently suffer from different biases. Table 4 presents our decisions about whether potential biases identified were judged to be additive or proportional, along with our rationale. Lack of adjustment for confounding has been shown to produce an additive bias [26]. We assumed that measurement error in exposure (D2 of ROBINS-E) was classical random measurement error and therefore was a proportional bias that moved the estimated effect towards the null [13, 27]. Missing data and selection of participants introduces collider stratification bias [28], which we judged to be an additive bias [29].

**Table 4:** Bias judgement of each domain for the applied case study.

| Study Design | Domains |  | Type of Bias | Justification |
| --- | --- | --- | --- | --- |
| *Randomized controlled trials* | D1 | Randomization process | Additive | Problems in the randomization process introduce a mixture of bias due to confounding and bias in selection of participants into the study (see also observational studies below) |
|  | D2 | Deviations from intended interventions | None | No RCTs had problems in this domain |
|  | D3 | Missing outcome data | Additive | Missingness in outcome can cause selection then this induces a bias that adds on to the truth [26] |
|  | D4 | Measurement of the outcome | None | No RCTs had problems in this domain |
|  | D5 | Selection of the reported result | Additive | We assume those who selectively report results are likely to favour a particular direction of effect |
| *Conventional observational studies of exposure effects* | D1 | Confounding | Additive | Confounding shifts effect estimates in a particular direction (rather than towards or away from the null) [26] |
|  | D2 | Measurement of the exposure | Proportional | If we assume classical measurement error in the exposure, this has a multiplicative effect on the estimate [27] |
|  | D3 | Selection of participants into the study (or into the analysis) | Additive | If both exposure and outcome are associated with selection into the study, then this induces a bias that adds on to the truth [26] |
|  | D4 | Post-exposure interventions | Additive | Only one study [56] had moderate bias, originally was a RCT changed to case-control due to lack of finding therefore suspect author is favouring a particular direction of effect |
|  | D5 | Missing data | Additive | If both exposure and outcome are associated with missingness then this induces a bias that adds on to the truth [26] |
|  | D6 | Measurement of outcomes | None | No observational studies had problems in this domain |
|  | D7 | Selection of the reported result | Additive | We assume authors who selectively report results are likely to favour a particular direction of effect |
| *Mendelian randomization* | D1 | Weak instrument | None | No MR studies had problems in this domain |
|  | D2 | Genetic confounding | Additive | Same as confounding |
|  | D3 | Other confounding | None | No MR studies had problems in this domain |
|  | D4 | Additional direct effects between instrumental variable and outcome (exclusion restriction assumption) | Additive | Based on Bowden *et al.* [57] this bias is an additive effect |
|  | D5 | Selection of participants | Additive | If both exposure and outcome are associated with selection into the study, then this induces a bias that adds on to the truth [26] |

Since our purpose here is illustrative, we used the default prior distributions (Table 5) in *triangulate* [14] to quantify the extent of bias (derived from [4]; see Discussion for further detail on the limitations of this approach, and potential solutions). Different prior distributions correspond to the different levels of risk-of-bias judgements. The default prior assumes that a high risk-of-bias equates to a large magnitude of bias, but with large variance so as to reduce the impact of that study on the pooled estimate. Using the default prior distributions for *triangulate* [14], we specified the prior distribution for “very high risk of bias” as double the value of mean and variance of the prior distribution for “high risk of bias”. For additive biases, the sign of the distribution is defined by the absolute direction of bias, with ‘favours lower exposure’ (“Favours comparator”) having a positive sign and ‘favours higher exposure’ (“Favours experimental”) having a negative sign (e.g. N(−0.09, 0.05)). As our effect measure is the risk ratio, the additive and proportional bias are applied to the estimate on the logarithmic RR scale.

**Table 5:** Prior distributions mapped to different extents of bias - Prior values for additive are defined as normal distributions, N(µ, σ^2^) with mean (µ) and variance (σ^2^), for proportional values are defined as log-normal distribution, Log-N(µ, σ^2^).

| Bias Level | Additive bias ("Favours experimental"/ "Favours comparator") | Proportional bias ("Towards null"/ "Away from null") | "Unpredictable" for additive bias | "Unpredictable" for proportional bias |
| --- | --- | --- | --- | --- |
| Low | - | - | - | - |
| Moderate | $N(0.09, 0.05)$ | $\text{Log-N}(0.03, 0.016)$ | $N(0, 0.05)$ | $\text{Log-N}(0, 0.016)$ |
| High | $N(0.18, 0.1)$ | $\text{Log-N}(0.06, 0.032)$ | $N(0, 0.1)$ | $\text{Log-N}(0, 0.032)$ |
| Very high | $N(0.36, 0.2)$ | $\text{Log-N}(0.12, 0.064)$ | $N(0, 0.2)$ | $\text{Log-N}(0, 0.064)$ |

### Combining relevant studies

We first estimated a combined unadjusted RR across the studies using random-effects meta-analysis of logarithmic (to base e) RRs, with a restricted maximum likelihood estimator to estimate between-studies heterogeneity. We used the *robviz* R package for meta-analysis and risk-of-bias visualization [30]. We used the RR for all incident (i.e. both fatal and non-fatal) CVD (or CHD) if a study provided both mortality and all incidences. We then obtained bias-adjusted effect estimates for each study and performed a random-effects meta-analysis with bias-adjusted effect estimates via *triangulate* [14]. All analyses were implemented in R version 4.2.2 [31].

To test sensitivity to these default prior distributions, we used an alternative set of priors (eTable 6), in which each level of risk of bias has the same mean bias but with different precision. This prior demonstrates that for high risk of bias, we are confident that bias exists, and therefore the interval is narrower, whereas for moderate bias, we are less confident about the existence of bias, hence larger variance. The prior for “unpredictable” bias remains the same as the default prior, as we are unsure of the bias direction.

## Results

We present the results for CHD; those for CVD were similar and presented in eAppendix 3. For brevity, we have not included all details of included studies here – we refer readers to the original systematic reviews for these [19–21].

### Description of studies

For the analysis of dietary beta-carotene on risk of CHD, there were three RCTs [32–34], seven MVR studies [35–39] and one MR study [21] (which provided three effect estimates from separate instrument-outcome data), shown in Table 6. Five studies defined their endpoint as patients experiencing any CHD events, one study with CHD mortality, and four studies with all MI events; three studies had separate numbers for all CHD and all MI events.

**Table 6:** Characteristics of included studies where endpoint is coronary heart disease (CHD)

| First author | Study design | Outcome* | Exposure summary | Median age (years)^ | Follow-up (years) | Relative risk* (95% CI) |
| --- | --- | --- | --- | --- | --- | --- |

| Randomized controlled trials |  |  |  |  |  |  |
| --- | --- | --- | --- | --- | --- | --- |
| Hennekens [32] | RCT (Physicians' Health Study) | All MI events | 50 mg/2d | 40-84 | 12.9 (Dur. 12) | 0.96 (0.85, 1.08) |
| Tornwall [33] | RCT (The Alpha-Tocopherol, Beta-Carotene Cancer Prevention Study) | All CHD events | 20 mg/d | 57 | 12.1 (Dur. 6.1) | 1.03 (0.92, 1.16) |
|  |  | All MI events |  |  |  | 1.07 (0.91, 1.24) |
| Cook [34] | RCT (Women's Antioxidant Cardiovascular Study) | All CHD events | 50 mg/2d | 60.6 | 9.4 (Dur. 9.4) | 1.00 (0.89, 1.13) |
|  |  | All MI events |  |  |  | 0.97 (0.77, 1.23) |
| Conventional observational studies |  |  |  |  |  |  |
| Osganian [35] | Cohort study (Nurses' Health Study) | All CHD events | Dietary | 50 | 12 | 0.83 (0.72, 0.96) |
| Todd [36] | Cohort study (Scottish Heart Health Study) | All CHD events | Dietary | 50 | 7.5 | 0.87 (0.68, 1.12) |
| Pandey [37] | Cohort study (Western Electric Company Study) | CHD mortality | Dietary | 40-55 | 21 | 0.84 (0.66, 1.09) |
| Klipstein-Grobusch [38] | Cohort study (The Rotterdam Study) | All MI events | Dietary | 67.45 | 4 | 0.22 (0.07, 0.68) |
| Koh [39] | Nested case-control study (Singapore Chinese Health Study) | All CHD events | Plasma | 69.2 (7.5) | 6.5 | 0.95 (0.66, 1.37) |
| Karppi [58] | Cohort study (Kuopio Ischaemic Heart Disease Risk Factor Study) | All MI events | Plasma | 57.1 | 11.5 | 0.64 (0.45, 0.93) |
| Hak [59] | Nested case-control study (Physicians' Health Study) | All MI events | Plasma | 58 | 6.3 | 0.82 (0.63, 1.07) |
| Mendelian randomization studies |  |  |  |  |  |  |
| Luo [21] | Mendelian randomization study | All CHD events | Plasma | Multiple GWAS~ | Multiple GWAS~ | 1.05 (0.95, 1.17)<sup>a</sup> |
|  |  |  |  |  |  | 1.02 (0.93, 1.11)<sup>b</sup> |
|  |  |  |  |  |  | 1.03 (0.84, 1.26)<sup>c</sup> |

CHD, coronary heart disease; MI, myocardial infarction; CI, confidence interval; Dur., intervention duration.
\*Relative risk is extracted from systematic reviews, not individual studies, except for the single MR study, extracted from Figure 2 of Luo *et al.*
^this varies between studies, some provides median, mean or range.
~Multiple GWAS: summary statistic for instrument-exposure is from the Nurses' Health Study, and instrument-outcome are from (a) Coronary Artery Disease Genome-Wide Replication and Meta-analysis plus the Coronary Artery Disease Genetics, (b) UK Biobank and (c) FinnGen study.

### Risk-of-bias

Results of risk-of-bias assessments for studies reporting CHD as an outcome are presented alongside the study results in Figure 2. Further detail of the risk-of-bias assessment is provided in Online Supplementary Tables. We judged two out of the three RCTs to have some concerns around bias, because of insufficient detail about randomization methods in bias domain 1 (D1). One RCT was judged to have a high risk of bias due to missing data (D3), and another had some concerns over the possibility of selection of the reported result (D5).

**Figure 2:**
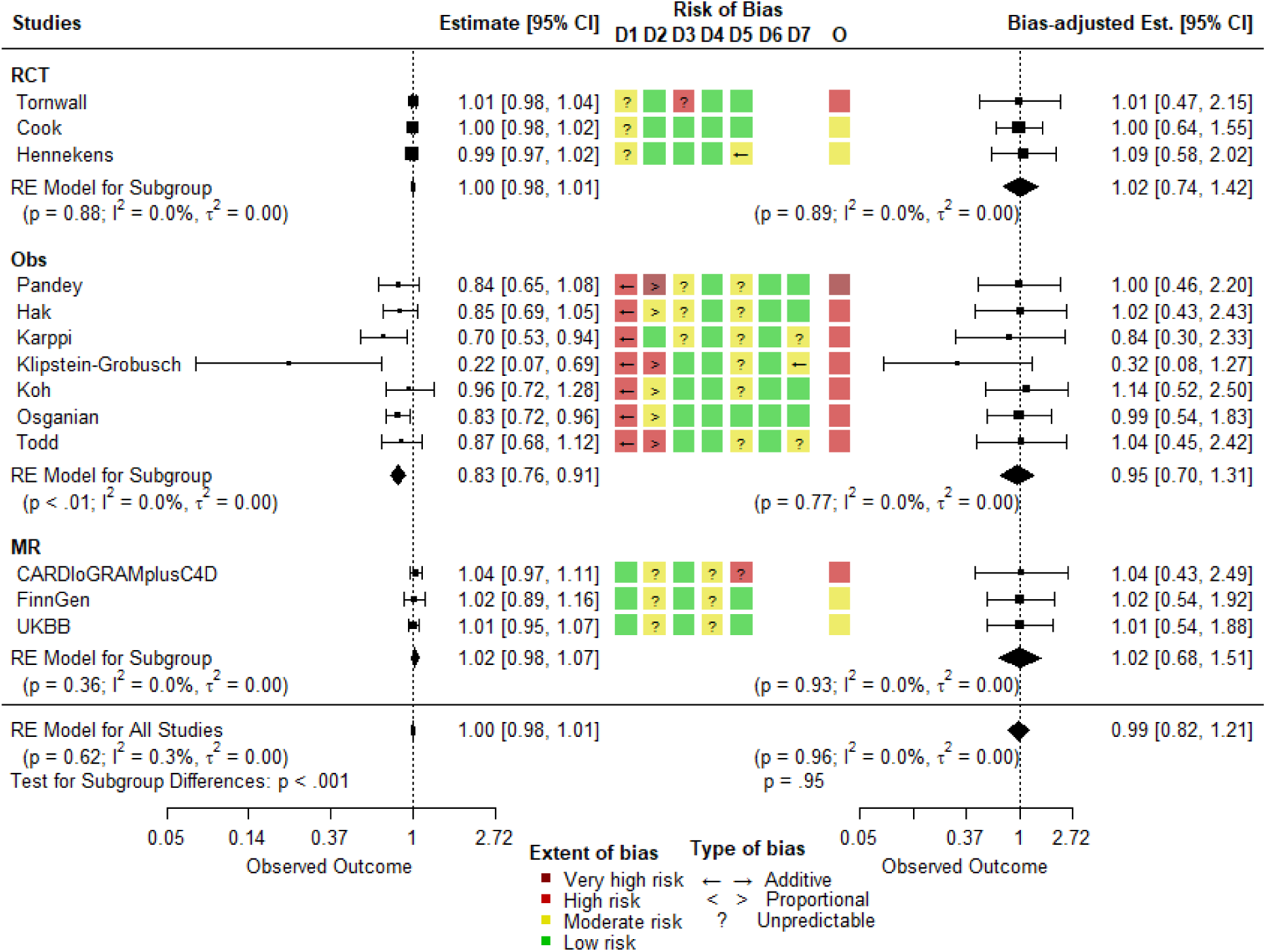
Unadjusted (left panel) and bias-adjusted (right panel) RR for random effects meta-analysis with CHD events as endpoint, and risk-of-bias assessment (middle panel). See *Table 2* for description of D1-D7 of each bias assessment tool. O is the overall judgment of the risk of bias. RCT, randomized controlled trial; Obs, conventional observational study; MR, Mendelian randomization; CI, confidence intervals; RE, random effects; p, p-value of the summary effect estimate; I^2^, heterogeneity variance / total variance; τ^2^, estimated amount of heterogeneity. The colours and symbols are shown in the legend on the plot.

The MVR studies were mainly judged to have high risk of bias overall. All the studies had adjusted for the minimal set of essential confounders (eTable 5), although still had high risk of bias due to confounding (D1) (as discussed above). We judged that residual confounding would result in additive bias in favour of increased beta-carotene intake [26]. Risk of bias from measurement of the exposure (D2) was considered high or very high for three of the studies. Although a food frequency questionnaire (FFQ) had been used to assess the exposure, we considered it unlikely that this would provide an accurate measure of an individual’s beta-carotene intake due to high day-to-day variability and poor recall [40]. We judged D2 to have proportional bias, as classical random measurement error is likely to pull the effect towards the null [13]. Pandey *et al.* [37] was the only MVR study to have a very high risk of bias overall, arising from measurement of exposure (D2) [37], since they did not implement the commonly used FFQ and beta-carotene was estimated from intake of other vitamins. Some studies were judged to have moderate risk of bias in selection of the reported result (D7) because of the lack of a pre-determined analysis plan.

All three of the MR analyses within one study have similar risk-of-bias judgements for each domain, apart from CARDIoGRAMplusC4D which had high risk of bias in D5. This is due to inclusion of a non-homogenous population for the instrument-outcome association when the instrument-exposure associations are estimated only using people of European descent. Therefore, the two different ancestries could confound the relationship between instrument and outcome via population stratification [2].

### Meta-analyses without bias adjustment

Meta-analysis of results unadjusted for bias from RCTs and MR studies provided no strong evidence of an effect of beta-carotene on CHD and did not show evidence of heterogeneity between studies (left panel of Figure 2). MVR studies gave a summary RR of 0.83 with a 95% CI from 0.76 to 0.91, with evidence of heterogeneity. The overall RR of three study designs was 1.00 (95% CI=0.98 to 1.01) and without evidence of heterogeneity. However, when differences between study designs were accounted for in meta-regression (labelled as “subgroup” in Figure 2), there is evidence of heterogeneity. Note that the estimates on the left-hand panel of Figure 2 differ from the estimates in Table 6, because the former have already been through the “map to common metric” step.

### Meta-analyses with bias-adjusted estimates

Meta-analysing the bias-adjusted effect estimates gave no strong evidence of an effect of beta-carotene on CHD from RCT, MVR, and MR studies (right panel of Figure 2). The summary RRs were 1.02 (95% CI=0.74 to 1.42), 0.95 (95% CI=0.70 to 1.31) and 1.02 (95% CI=0.68 to 1.51) from RCTs, MVR and MR analyses, respectively (Figure 2). The overall RR for all studies was 0.99 (95% CI=0.82 to 1.21), and without evidence of heterogeneity between three study designs. There was one RCT and one MR analysis with high risk of bias, but we judged the direction of bias to be unpredictable in both cases, so that the adjusted effect estimate was unchanged by inclusion of these studies (but with a wider CI).

With a different set of priors (eTable 6), similar results were obtained in the sensitivity analysis (eFigure 1): the summary RR is 1.01 (95% CI=0.82 to 1.23) and without evidence of heterogeneity between the study designs (right panel of eFigure 1).

## Discussion

Our focus on methods for bias quantification and adjustment is an extension of the triangulation approach initially described by Lawlor *et al.* [1]. Our framework identifies the target question and the relevance of each study to this and uses formal risk-of-bias tools to identify different sources of internal bias. We have shown how to address the potential biases by stating the expected magnitude and direction of bias for each study through informative prior distributions for internal bias. Our illustrative example demonstrates how bias adjustment can be applied in triangulation. In this example, even with the default prior distributions, the bias-adjusted results across our three study types were homogeneous, suggesting that bias-adjustment has removed much of the variability between study estimates and thus increased our confidence in the overall conclusion (which in this case is that there is no evidence that dietary intake of beta-carotene affects risk of any CVD or of CHD). Furthermore, the same conclusion was replicated with a different prior distribution. The detailed risk-of-bias assessments did not predict opposing net directions of bias across the included study designs. However, since the assessments did not suggest that the net biases across studies operated in a single direction (being either null or towards a positive effect of beta-carotene), we proceeded with the quantitative triangulation analysis to illustrate the approach.

In our example, bias adjustment did not change the overall estimate materially, although did increase the width of the confidence interval around it due to our uncertainty in the biases. If there is less heterogeneity after bias adjustment, as here, this increases confidence in the result (which in this case is a lack of evidence against the null hypothesis). A reduction in heterogeneity is the expected outcome of a successful triangulation exercise. If bias adjustments are applied when all biases are believed to be in the same direction (i.e. bias-adjustment outside of a triangulation context), we would not expect to see as much reduction in between-study heterogeneity after adjustment and we would not generate the reassurance that can be obtained from a triangulation exercise.

It is common practice in meta-analysis to restrict the analysis to studies considered to be most relevant and at lowest risk of bias (although sensitivity analyses might be conducted to evaluate the influence of high risk of bias results). We have exemplified here through the use of prior distributions for bias, which is one approach to quantifying bias. Expert opinion to inform such prior distributions [4, 41, 42] can come in different forms: different choices of bias terms via sensitivity analysis [43], distributions of bias terms [4] or modelling “ignorance” about the bias term [44]. Our bias adjustment method [4] makes two important assumptions. First, the bias distributions are assumed independent between domains. In practice, this may not be the case [4]. For example, the bias from measurement of exposure could be positively correlated with bias due to missing data, because dietary beta-carotene is measured from FFQ, which is time-consuming if it has many food items, and the participants are less likely to answer all the questions thoroughly. Second, the method proposed by Turner *et al.* [4] adjusts the effect estimate initially for additive bias and then proportional bias. However, the ordering of bias adjustment should reflect the order in which the biases operate, and so the order of adjustment should be considered for each study [45]. It is important to assess each bias domain in the context of each individual study. For example, here we considered that measurement error in estimating the dietary intake of beta-carotene in observational studies was likely to be random and unrelated to the outcome, thus biasing results towards the null. However, in different contexts, measurement error could have no effect on bias or bias away from the null, or the direction could be unclear.

For the sake of illustration, we used the default prior distributions from the R package *triangulate* [14], which represent expert opinion elicited from a study examining the effectiveness of routine anti-D prophylaxis in Rhesus negative women [4]. We see no particular reason why these distributions should not generalize to other causal questions. These default choices simplify the options to one prior distribution for each risk-of-bias judgement level (e.g. ‘moderate’, ‘high’ or ‘very high’ risk of bias), assume these distributions to be the same irrespective of the bias domain or the study design, and assume they are identical across studies. However, meta-epidemiologic studies of RCTs suggest that the effect of bias may be different for different bias domains [46], and further evidence is needed for studies other than RCTs [47]. The bias adjustment framework can allow prior distributions to be specific to each study result. Thus, a more complete analysis could use the risk-of-bias evaluations could be used to tailor the priors to each domain and each study. Lastly, future work should separate risk of bias from magnitude of bias, which are currently conflated by the default priors.

Specifying suitable prior distributions for bias is challenging. However, we believe it is often preferable to the alternatives of (i) making the very strong assumption that all estimates are unbiased (a prior expectation of zero bias with no uncertainty) and (ii) excluding studies altogether (a prior weight of 0). The aim of quantitative triangulation is to compare results of studies with differing predicted directions of bias – therefore, to carry out the triangulation exercise, we must be able to assess the risk or magnitude of bias to some degree. In the unlikely event that little was known about the risk or magnitude of bias for all (or most) studies, triangulation would not be appropriate. It can be difficult to decide how much heterogeneity in direction of bias justifies quantitative triangulation, versus meta-analysing only the high-quality studies (as is often done in meta-analyses). Future work could usefully explore how this judgement could be made quantitatively, for example building on standard heterogeneity statistics from the meta-analysis toolkit.

Our experience suggests that careful *a priori* definition of the research question (e.g. the causal effect of interest) is vital when embarking on a triangulation exercise. In our example, most of the primary studies did not clearly define their research question and rarely covered the six domains we used to characterize the target question. Consequently, we had to deduce the research questions from the study description. In future, we advocate specification of clearly defined causal questions in epidemiologic studies of the effects of exposures, as argued by Munafò and Davey Smith [48].

Judging the relevance of different studies to the target question is potentially more subjective than judging risk of bias. For some observed between-study differences, quantitative approaches can be used. For example, we included studies that measured circulatory beta-carotene and used correlation estimated from a large meta-analysis of dietary and plasma beta-carotene [25] to convert the RR for circulatory to dietary beta-carotene, assuming that this conversion was the same for everyone. The use of random-effects meta-analysis should capture variation between study populations (effect heterogeneity). Other approaches to tackle relevance include data fusion [49] and causal meta-analysis [50]. Both of these focus on transportability [51], estimating a causal effect for a target population by transporting causal effects from studies with varying distributions of effect modifiers [52]. However, these approaches only address one aspect of relevance, and do not consider bias separately – they assume that the main source of heterogeneity is differences between study populations. Furthermore, both approaches require individual-level data, which is seldom available for all studies. The *triangulate* R package [14] can express relevance judgements through a further set of prior distributions in the same way as for biases, addressing the three ‘relevance domains’ of population (or participants), exposure, and outcome (or endpoint), thereby allowing expression of uncertainty around relevance [11]. This could allow uncertainty in the conversion of the RR from circulating beta-carotene to an RR for dietary beta-carotene – whereas in our analyses we assumed that beta-carotene absorption does not vary with individual characteristics (e.g. sex, dietary habits). Further work is needed to establish guidance for assessing relevance.

Our case study highlights the need for a more thorough risk-of-bias tool for MR. The risk-of-bias tool for MR by Mamluk *et al.* [10] was designed for their own systematic review, unlike RoB 2 and ROBINS-E which are products of a team of multiple experts and many years of piloting. The tool by Mamluk *et al.* [10] also does not include domains for measurement error, sensitivity analysis or missing data [53].

All the studies in our illustrative example aligned in their conclusions after bias adjustment. In many cases, however, there may remain heterogeneity in the bias-adjusted estimates. As with any meta-analysis, the reasons for this should be examined, including considering whether the heterogeneity is the result of study-level variables. For example, we assumed sex does not moderate the beta-carotene-CVD relationship, an assumption which could be examined using meta-regression if there were enough studies with different proportions of men and women, or within a large study if individual-level data were available. In general, heterogeneity between effect estimates would give rise to further investigation, as it clearly indicates that the effect of interest cannot be reliably estimated from the evidence available.

Our applied example was an illustration of methodology and we assumed that the two source systematic reviews had followed best practices [54]. However, the reviews were published in 2018 and therefore will not include new evidence. We recommend the triangulation efforts should use evidence synthesis best practices, including comprehensive systematic or umbrella reviews to identify all relevant studies. Our example integrates evidence from three methods, and it would be valuable to explore how this framework expands to include other study designs, such as within-sibling and other family-based analyses, or cross context comparisons.

In conclusion, triangulation is a challenging and still-developing area in epidemiology. We have developed a framework for quantitative triangulation, including guidance for the process of identifying and addressing biases and differences in relevance, and illustrated how prior distributions can empirically lay out researchers’ assumptions about these factors. Our framework should be viewed as a tool for structuring and transparently documenting expert judgements regarding relevance and bias, rather than as a replacement for expert judgement.

## Acknowledgments

We would like to thank an anonymous reviewer and Jeremy Labrecque for insightful comments to help improve our paper.

## Author Contributions

All authors contributed to the study conception and design. Material preparation, data collection and analysis were performed by CYS. CYS, JAB, MCB, AGS, KT and JPTH were the assessors for risk of bias. LAM and CYS worked on software. The first draft of the manuscript was written by CYS, and KT and JPTH revised the manuscript critically for important intellectual content. All authors commented on previous versions of the manuscript. All authors read and approved the final manuscript.

## Ethics approval

No ethics approval required as only summary statistics were used.

## Data and Software code

Data are extracted from published work. Code is available at *CYShapland/BetaCarotene_CVD (github.com)*.

