## Supplementary Materials for "A quantitative approach to evidence triangulation: development of a framework to address rigour and relevance"

### Supplementary material for “A quantitative approach to evidence triangulation: development of a framework to address rigour and relevance”

### Appendix 1. Method

#### Appendix 1a. Relevance: assessing and mapping results to a comparable metric

*eTable 1: Considerations for study populations, exposure, exposure window and outcomes of the target causal question. Note that this list some of the considerations and is not an exhaustive list.*

| Domain | Considerations |
|---|---|
| Target Population | Is the effect in the study population expected to be plausibly the same as the effect in the target population? This would not be true if effect modification is expected.<br>Are age and/or sex effect modifiers?<br>Disease free at baseline?<br>Cross context analysis? Countries can vary by social norms, diet, etc. |
| Exposure Measure | How is the exposure measured? |
| Exposure Window | The exposure window of interest is the exposure time period for which the effect of the exposure on the outcome is being estimated. Specification should include both the time of onset and period of exposure. Example exposure windows include: lifetime exposure (from birth or from conception); exposure during ages 50-55 years; the period from first employment in a particular occupation; during childhood (birth to age 10 years), or during pregnancy.<br>If target duration is intake over 10 years, would an randomized control trial (RCT) lasting for 6 months be relevant?<br>For Mendelian randomization (MR) studies, it is assumed that the genetic instrument acts on the exposure from conception/foetal development onwards [1]. This assumption could be changed if there are different genetic instruments for different periods of exposure, as in a recent study of the effects of childhood vs adulthood body mass index (BMI) [2]. |
| Exposure over time should be summarized | Examples of how exposure might be summarized over time include: ever/never exposed; cumulative exposure; average exposure; or peak exposure during the exposure period. Alternatively, there may be only a single exposure event, or the exposure may be time invariant (such as a genetic variant or family history). |
| Outcome Measure | How is the outcome measured?<br>How long is the follow-up? Are individuals more likely to experience an event if the follow-up is for a long period of time? |
| Statistical Parameter | Statistical model used to relate outcome to exposure and time - e.g. logistic (estimate odds ratio), survival (estimate hazard ratios , linear regression (estimate additive regression coefficient), etc.<br>Per what unit increase in exposure is the effect on the outcome to be expressed, and is it assuming the effect on outcome of one unit increase in exposure is additive, multiplicative or following a threshold model, etc. |

##### *Appendix 1b. Bias adjustment*

Bias from confounding, the relationship of exposure (X), outcome (Y) and confounder (U) for individual *i* is;

$$Y_i = \beta_0 + \beta_1 X_i + \beta_3 U_i + \epsilon_i$$

$$X_i = \alpha_0 + \alpha_1 U_i + \epsilon_i$$

Let $\widehat{\beta_1}$ be the regression coefficient without adjusting for U.

The total additive bias for study *i* is defined as:

$$\mu_{i\delta} = \sum_j \alpha_{ij\delta},$$

$$\sigma_{i\delta} = \sum_j \psi_{ij\delta}^2,$$

where $\alpha_{ij\delta}$ and $\psi_{ij\delta}^2$ are the mean and variance values respectively from the prior distributions for additive bias *j* in the *i*th study.

The total proportional bias for *i*th study is defined as:

$$\mu_{i\beta} = \exp(\alpha_{i\beta} + \frac{\psi_{i\beta}^2}{2}),$$

$$\sigma_{i\beta} = \left(\exp(\psi_{i\beta}^2) - 1\right)\exp\left(2\alpha_{i\beta} + \psi_{i\beta}^2\right),$$

where $\alpha_{i\beta} = \sum_p \alpha_{ip\beta}$ and $\psi_{i\beta}^2 = \sum_p \psi_{ip\beta}^2$, $\alpha_{ip\beta}$ and $\psi_{ip\beta}^2$ denoting the mean and variance values, respectively, from the priors for the logarithm of proportional bias *p* in the *i*th study.

Then the bias adjusted point estimate is:

$$\widehat{\theta_\iota} = \frac{y_i - \mu_{i\delta}}{\mu_{i\beta}},$$

where $y_i$ is the log(RR) for *i*th study. The bias adjusted standard error is then:

$$SE(\widehat{\theta_\iota}) = \left(\frac{1}{\mu_{i\beta}}\right)^2 (s_i^2 + \sigma_{i\beta}^2 \widehat{\theta_\iota}^2 + \sigma_{i\delta}^2),$$

where $s_i$ is the standard error of log(RR) for the *i*th study. Note that these two equations are the same as Equation (6) and (7) in Turner *et al.* [3] but the parameters for indirectness are removed.

#### Appendix 2. Application to case-study

##### *Appendix 2a. Identification of relevant studies*

In this section we give a brief summary of the two systematic reviews and Mendelian randomization (MR) study, where we extracted the effect estimates for dietary (and circulating) beta-carotene, and cardiovascular disease (CVD) (and coronary heart disease (CHD)).

For conventional observational studies we identified a review [4] from 2018 which concluded from a meta-analysis of 69 prospective studies (from total of 46,082 identified) that higher dietary intake and/or blood concentrations of antioxidants was associated with reduced risk of CVD, total cancer, and all-cause mortality. The authors [4] included prospective cohort and nested case-control studies that reported adjusted risk ratios (RRs) of the association between vitamin C, vitamin E, and carotenoids in the diet or measured in blood and the risk of CHD, stroke, CVD, total cancer, and all-cause mortality. They excluded studies that only assessed supplemental intake of these antioxidants and studies that did not adjust for any confounders (the authors did not specify a list of confounders). The review reported a RR from each study which had undergone a series of data transformations before being meta-analysed. Studies with multiple RRs for categories of beta-carotene were converted into a single RR [5] and the number of cases and non-cases were imputed for studies that did not report them. We have used the transformed RRs in our meta-analysis.

A systematic review from 2018 of randomized control trials (RCTs) for vitamin and mineral supplements and CVD and all-cause mortality [6] identified 179 studies that fulfilled their inclusion criteria (from 1,496 studies) and these were included in a meta-analysis. The review excluded studies that had no control arm and where intervention duration was less than 6 months. From each study, the authors extracted the number of cases and total participant numbers for supplement/intervention and control groups, and obtained the summary statistics based on these numbers using the Mantel-Haenszel method [6].

We identified one MR study [7] from 2021. It included 768,121 participants with 93,230 who had CHD and examined the effect of circulating beta-carotene on CHD risk [7]. The authors performed two-sample MR, with single nucleotide polymorphisms (SNPs) selected for use as instrumental variables from the Nurse's Health Study based on SNPs that were genome-wide associated ($p < 5 \times 10^{-8}$) and independent of each other (linkage disequilibrium (LD) threshold of $r^2 < 0.2$). The authors extracted the summary results of the association of these SNPs with beta-carotene from the same study and for the association of the same SNPs with CHD (outcome) from the CARDIoGRAMplusC4D consortium, the UK Biobank and the FinnGen Study with causal effect estimates of circulating beta-carotene on CHD from the three studies combined using fixed effects meta-analysis. We treat the three estimates as arising from three distinct studies as they have used three different populations for the SNP-outcome association.

#### *Appendix 2b. Assessing relevance*

***eTable 2:*** *Population and exposure for included studies where outcome is coronary heart disease (CHD)*

| *First author* | *Country* | *Population* | *Exposure measure* | *Age range (years)^* | *Follow-up (years)* | *Outcome* |
|---|---|---|---|---|---|---|
| *Randomized controlled trials* | | | | | | |
| Hennekens [11] | US | Physicians | 50 mg/2d | 40-84 | 12.9 (Dur. 12) | All myocardial infarction (MI) events |
| Tornwall [24] | Finland | Smokers | 20 mg/d | 50-69 | 12.1 (Dur. 6.1) | All CHD events |
| | | | | | | All MI events |
| Cook [12] | US | Female patients with history of CVD | 50 mg/2d | ≥40 | 9.4 (Dur. 9.4) | All CHD events |
| | | | | | | All MI events |
| *Conventional observational Studies* | | | | | | |
| Osganian [25] | US | Nurses | Dietary | 38-63 | 12 | All CHD events |

| | | | | | | |
|---|---|---|---|---|---|---|
| Todd [26] | Scotland | General population | Dietary | 40-59 | 7.5 | All CHD events |
| Pandey [27] | US | General population | Dietary | 40-55 | 21 | CHD mortality |
| Klipstein-Grobusch [28] | Netherlands | Older population | Dietary | 55-95 | 4 | All MI events |
| Koh [29] | Singapore | General population | Plasma | 45-74 | 6.5 | All CHD events |
| Karppi [30] | Finland | General population | Plasma | 46-65 | 11.5 | All MI events |
| Hak [31] | US | Physicians | Plasma | 40-84 | 6.3 | All MI events |
| *Mendelian randomization studies* | | | | | | |
| Luo [7] | US, UK and Finland | Nurses and general population | Plasma | Multiple GWAS~ | Multiple GWAS~ | All CHD events |

***eTable 3:*** *Population and exposure for included studies where outcome is cardiovascular disease (CVD)*

| *First author* | *Country* | *Population* | *Age range (years)* | *Exposure measure* | *Follow-up (years)* | *Outcome* |
|---|---|---|---|---|---|---|
| *Randomized controlled trials* | | | | | | |
| Greenberg [8] | US | Biopsy-proved basal cell or skin cancer patients | 27-84 | 50 mg/d | 8.2 (Dur. 4.3) | CVD mortality |
| Green [9] | Australia | Healthy and patients who previously had skin cancer | 40-84 | 30 mg/d | 4.5 (Dur. 4.5) | CVD mortality |
| Lee [10] | US | Females in general population | 20-69 | 50 mg/2d | 4.1 (Dur. 2.1) | All CVD events |
| Hennekens [11] | US | Physicians | 54.6* (SD=7) | 50 mg/2d | 12.9 (Dur. 12) | All CVD events |
| | | | | | | CVD mortality |
| Cook [12] | US | Female patients with history of CVD | 27-84 | 50 mg/2d | 9.4 (Dur. 9.4) | All CVD events |
| | | | | | | CVD mortality |
| *Conventional observational Studies* | | | | | | |
| Genkinger [13] | US | General population | 30-93 | Dietary | 12.2 | CVD mortality |
| Stepaniak [14] | Europe | General population | 45-69 | Dietary | 7.23 | CVD mortality |
| Buijsse [15] | Netherlands | Older population | 65-84 | Dietary | 15 | CVD mortality |
| de Oliveira Otto [16] | US | Patients with subclinical CVD | 45-84 | Dietary | 6.2 | All CVD events |
| Fletcher [17] | UK | Older population | 75–84 | Plasma | 4.4 | CVD mortality |
| Bates [18] | UK | General population | 65-99 | Plasma | 13.5 | CVD mortality |

| | | | | | | |
|---|---|---|---|---|---|---|
| Goyal [19] | US | General population | >20 | Plasma | 14.2 | CVD mortality |
| Greenberg [8] | US | Biopsy-proved basal cell or squamous cell skin cancer patients | 27-84 | Plasma | 8.2 | CVD mortality |
| Kilander [20] | Sweden | General population | 48.6-51.2 | Plasma | 25.7 | CVD mortality |
| Karppi [21] | Finland | General population | 46-65 | Plasma | 15.9 | CVD mortality |
| Mezzetti [22] | Italy | older population | >80 | Plasma | 3.95 | All CVD events |
| Sesso [23] | US | Physicians | 69.7* (SD= 8.1) | Plasma | 2.1 | All CVD events |

*Mean and standard deviation (SD) available in study.

#### Exposure window

We considered various possible exposure windows of interest, including: from birth; from age 40 years; from 5 years before cohort recruitment; or from time of cohort recruitment. Different choices may be expected to result in different types of bias being identified (eTable 4). For example, choosing the exposure window of interest to be from birth would minimize the risk of baseline confounding by prior beta-carotene intake (because there is no dietary intake before the start of the exposure window), but would be associated with very substantial selection bias (because the sample is usually not identified until many years after the start of the exposure window – e.g., individuals could have experienced an event). On the other hand, choosing the exposure window of interest to be from the time of recruitment into the study would lead to a high risk of bias due to confounding by previous beta-carotene intake (because intake at baseline is likely to reflect very strongly a person's intake in recent years), but would minimize selection bias (because there is minimal opportunity for participants to be selected out of the study after the start of the exposure window).

***eTable 4**: Points of consideration when specifying exposure window for evaluation of conventional observational (cohort) studies of the effect of beta-carotene on cardiovascular disease or coronary heart disease in middle aged adults*

| **Exposure window** | **Domains** | **Assessment of bias** |
|---|---|---|
| From conception/foetal development | *Confounding by previous exposure* | Low risk of bias |
| | *Exposure measurement* | High risk of bias because middle-aged exposure may be a poor representation of lifetime exposure |
| | *Selection into the study* | Very high risk of bias because many individuals will be missing (died or had CVD/CHD before middle age) |
| *From age 40 years** | *Confounding by previous exposure* | High risk of bias because exposure level at time of recruitment will reflect recent exposure (and recent exposure may be predictive of CVD/CHD) |
| | *Exposure measurement* | Probably low risk of bias, assuming exposure levels are reasonably consistent during middle to older age |
| | *Selection into the study* | High risk of bias in studies that recruit people older than 40, because some individuals will be missing (died or had CVD/CHD between age 40 and study recruitment) |

| | | |
|---|---|---|
| From 10* years before cohort recruitment/exposure measurement | *Confounding by previous exposure* | High risk of bias because exposure level at time of recruitment will reflect recent exposure (and recent exposure may be predictive of CVD/CHD) |
| | *Exposure measurement* | Probably low risk of bias, assuming exposure levels are reasonably consistent during middle to old age |
| | *Selection into the study* | Problematic, because some individuals will be missing (died or had CVD/CHD in the 10 years before study recruitment) |
| From the time of cohort recruitment/exposure measurement | *Confounding by previous exposure* | Very high risk of bias because exposure level at time of NRSE recruitment will strongly reflect recent exposure (and recent exposure may be predictive of CVD/CHD). |
| | *Exposure measurement* | Low risk of bias, because measured directly in the study |
| | *Selection into the study* | Low risk of bias, since participants should not be lost between start of exposure and recruitment to the study |

** Exemplar timing – other values could be used; NRSE, non-randomized study exposure*

#### *Appendix 2c. Map to common metric*

##### Conversion factor (k) from circulating to dietary beta-carotene

A conversion factor between the average change in dietary beta-carotene (y) for every unit increase in circulating beta-carotene (x) is given by

$$k = r_{xy}\,\frac{\sigma_y}{\sigma_x}$$

where $\sigma_y$ and $\sigma_x$ are the standard deviations of dietary and circulating beta-carotene, respectively, and $r_{xy}$ is the correlation between them. We found a meta-analysis from 2015, of 53 studies that estimated the correlation between dietary and plasma beta-carotene [32], in which we extracted $r_{xy}$= 0.27, weighted means (from meta-analysis) of dietary beta-carotene intake of 3924.7 µg/day (95% CI 3383.8 to 4465.5, n=67) and of circulating beta-carotene of 0.47 µmol/L (95% CI 0.46 to 0.48, n=78). We used the CI width and sample size to estimate the standard deviations of dietary and circulating beta-carotene levels, in units of µg/d and µmol/L, respectively.

##### Conventional observational Study

RRs for circulating beta-carotene in the systematic review of observational studies were given per 25 µg/dL.

*Step 1*: Calculate conversion factor (k) to convert circulating beta-carotene (µg/dL) to dietary beta-carotene (µg/d). RRs for circulating beta-carotene were given per 25 µg/dL rather than per µmol/L. With the values from section "Conversion factor (k) from circulating to dietary beta-carotene" and using 0.01863 to convert µmol/L to µg/dL as Burrows *et al.*'s circulating beta-carotene was in µmol/L therefore k is:

$$k = r_{xy}\,\frac{\sigma_y}{\sigma_x}$$

$$= 0.27 \times \frac{\sqrt{67} \times \frac{4465.5 - 3383.8}{3.92}}{\sqrt{78} \times \frac{(0.48 - 0.46)/0.01863}{3.92}}$$

$$= 252.14$$

*Step 2*: To harmonize all the RR estimates to be on the same unit and dosage, per 5,000 µg/day, we therefore converted as follows:

$$\log(RR_{Diet}) = \frac{5000}{k(= 252.14) \times 25} \times \log(RR_{Circ}),$$

where $RR_{Diet}$ and $RR_{Circ}$ are the risk ratios for dietary and circulating beta-carotene, respectively.

#### Mendelian Randomization Study

The effect sizes from MR studies were per µg/L in log-transformed circulating beta-carotene.

*Step1*: To transform the effect size approximately to per µg/L untransformed circulating beta-carotene (estimated at the study average). We assume the fitted model is

$$\hat{Y} = \hat{\alpha} + \hat{\beta} \times \log(X)$$

We assume X and Y are linearly related. Then using the Taylor expansion of log(X) at the overall mean of X, E[X] gives

$$\ln(E[X]) + \frac{1}{E[X]}(X - E[X]) - \cdots.$$

We assume 2nd, 3rd term and etc of the Taylor expansion is small and can be omitted. Substitute X into the fitted model:

$$\begin{aligned}\hat{Y} &= \hat{\alpha} + \hat{\beta} \times \left[\ln(E[X]) + \frac{1}{E[X]}(X - E[X])\right] \\ &= \hat{\alpha} + \hat{\beta} \times \ln(E[X]) + \frac{\hat{\beta}}{E[X]}(X - E[X]) \\ &= \hat{\alpha} + \hat{\beta}[\ln(E[X]) - 1] + \frac{\hat{\beta}}{E[X]}X\end{aligned}$$

Hence the coefficient for untransformed X is $\frac{\hat{\beta}}{E[X]}$.

*Step 2*: Calculate conversion factor (k) to convert circulating beta-carotene (µg/L) to dietary beta-carotene (µg/d). With the values from section "Conversion factor (k) from circulating to dietary beta-carotene", 0.01863 to convert µmol/L to µg/dL and 10 to convert µg/dL to µg/L, k is:

$$\begin{aligned}k &= r_{xy}\,\frac{\sigma_y}{\sigma_x} \\ &= 0.27 \times \frac{\sqrt{67} \times \frac{4465.5 - 3383.8}{3.92}}{\sqrt{78} \times \frac{(((0.48 - 0.46) \times 10)/0.01863}{3.92}} \\ &= 25.21\end{aligned}$$

*Step 3*: Combine the information from Step 1 and Step 2, we converted circulating beta-carotene to per 5000 µg/d dietary beta-carotene using:

$$\log(RR_{Diet}) = \frac{5000}{k\ (=25.21)} \times \frac{\log(RR_{Circ})}{\mu_y},$$

where $\mu_y$ is the mean of the circulating beta-carotene (µg/L) from the MR study.

##### Randomized control trial

For each RCT, we converted RR for controls vs beta-carotene supplements to per 5,000 µg/day, by dividing logarithmic RR by dosage multiplied by 5,000 [33].

#### *Appendix 2d. List of Confounders*

***eTable 5**: List of confounders*

| **Confounders** | **Related variables** |
|---|---|
| **Minimal** | |
| Age | |
| Sex | |
| SES | Socioeconomic position (SEP), socioeconomic status (SES), family income, housing tenure and education. |
| Ethnicity | |
| BMI | |
| Dietary intake | fruit & vegetables intake, total energy intake and supplement intake |
| **Desirables** | |
| Family history of CVD | |
| Comorbidities | Cancer and diabetes. |
| Blood pressure | |
| Smoking | |
| Alcohol consumption | |
| Physical activities | |
| Circulating lipid levels | |
| Circulating vitamins | |

#### *Appendix 2e. Prior sensitivity*

***eTable 6:** Prior sensitivity - Prior distributions mapped to different extents of bias - Prior values for additive are defined as normal distributions, N(μ, $\sigma^2$) with mean (μ) and variance ($\sigma^2$), for proportional values are defined as log-normal distribution, Log-N(μ, $\sigma^2$).*

| Bias Level | Additive bias ("Favours experimental"/ "Favours comparator") | Proportional bias ("Towards null"/ "Away from null") | "Unpredictable" for additive bias | "Unpredictable" for proportional bias |
|---|---|---|---|---|
| Low | - | - | - | - |
| Moderate | N(0.18, 0.2) | LogN(0.06, 0.064) | N(0, 0.05) | Log-N(0, 0.016) |
| High | N(0.18, 0.1) | LogN(0.06, 0.032) | N(0, 0.1) | Log-N(0, 0.032) |
| Very high | N(0.18, 0.05) | LogN(0.06, 0.016) | N(0, 0.2) | Log-N(0, 0.064) |

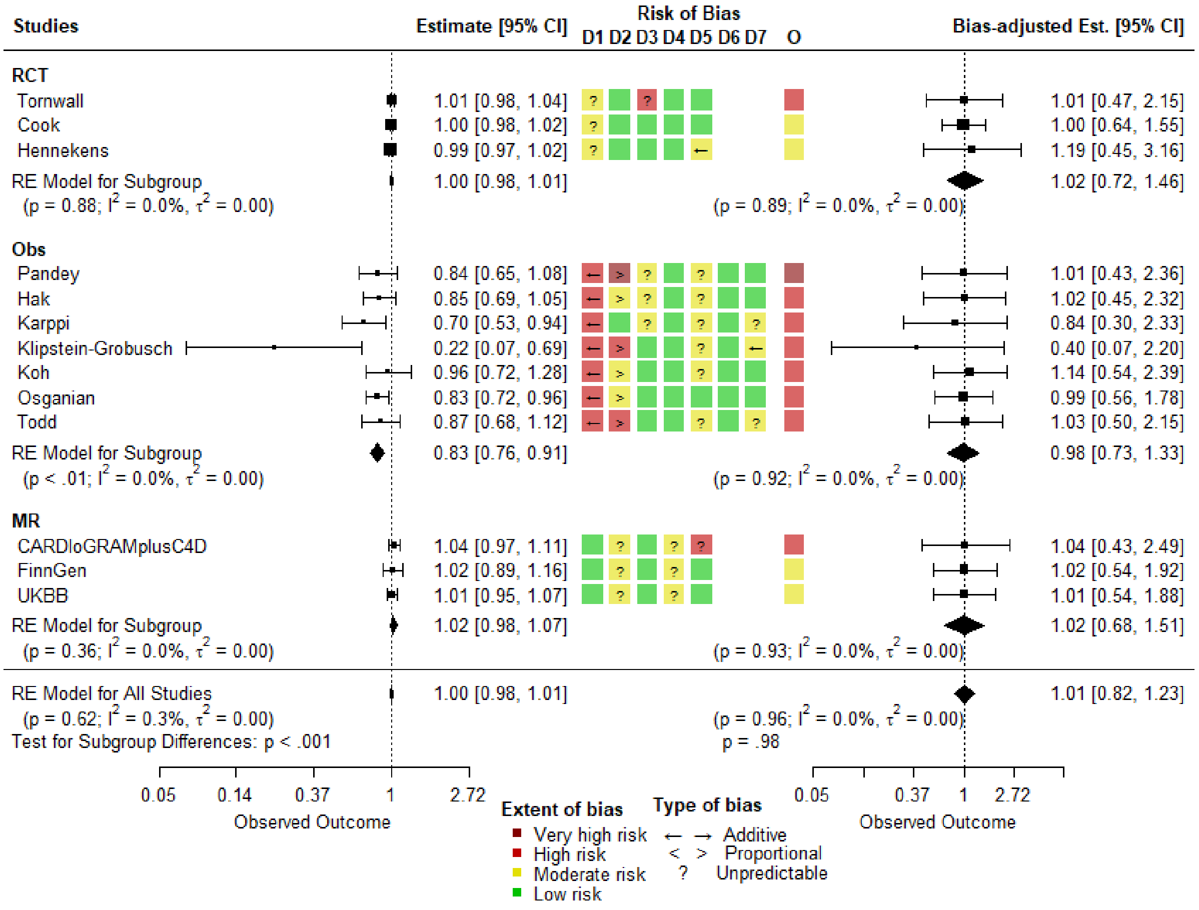

***eFigure 1**: Prior Sensitivity-Unadjusted (left panel) and bias-adjusted (right panel) random effects meta-analysis for CHD events as endpoint with risk-of-bias assessment (middle panel). See Table 1 for description of D1-D7 of each bias assessment tool. Priors is defined in eTable 6. O is the overall judgment of risk of bias. RCT, randomized controlled trial; Obs, conventional observational study; CI, confidence intervals; p, p-value of the pooled effect estimate; $I^2$, heterogeneity variance / total variance; $\tau^2$, estimated amount of heterogeneity. The colours and symbols are shown in the legend on the plot.*

#### *Appendix 3. results for cardiovascular disease (CVD)*

##### Description of studies

A total of 17 studies [8-23,34] were included in the analysis of dietary beta-carotene intake on the risk of CVD (the broad outcome; CHD is considered separately below); five were RCTs and 12 were MVR studies (**eTable 6**). There were no MR studies on the relationship between beta-carotene and CVD. Only four MVR studies measured dietary beta-carotene, with the rest measuring circulating levels. Most of the studies defined their endpoint as CVD mortality; five studies included non-fatal CVD events with CVD mortality (referred to as all CVD events), and two studies presented separate effect sizes for all CVD events and for CVD mortality.

##### Risk-of-bias

Results of risk-of-bias assessments for studies reporting CVD as an outcome are presented alongside the study results in the middle panel of **eFigure 1**. We judged for all of the RCTs to have “moderate” risk of bias, either because of insufficient detail about randomization methods or the possibility of selection of the reported result. Most of the MVR studies had adjusted for what we a priori defined as the minimum set of essential confounders (**eTable 5**) only, and so were judged to be at high risk of bias for confounding (D1). We judged that not adjusting for confounding, the bias again predicted to favour a beneficial effect of beta-carotene. Risk of bias from measurement of the exposure (D2) was considered high for half of the studies. Unlike other studies, the effect estimate of Genkinger *et al.*[13] is above 1, hence proportional bias is pointing left rather than right to pull towards the null. Buijsse *et al.* [15] had a very high risk of bias arising from D2 because the authors did not use the commonly used FFQ (their FFQ was adapted from [36]).

#### Meta-analyses without bias correction

The meta-analysis of RCTs provided little evidence of an effect of beta-carotene on risk of CVD, with a RR of 1.01 (95% CI=0.99 to 1.02) (left panel of **eFigure 1**), and little heterogeneity between studies ($I^2$=3.6%, $\tau^2$=0). The meta-analysis of the MVR studies indicated a protective effect of beta-carotene against CVD (RR=0.91, 95% CI=0.85 to 0.97), but with greater between-studies heterogeneity ($I^2$=18.7%, $\tau^2$=0). The summary RR for all studies was 1.00 (95% CI=0.99 to 1.01) and evidence for heterogeneity between study designs (p-value=0.003).

#### Meta-analysis with bias corrected estimates

The right-hand plot of **eFigure 1** gives the impact of correcting for bias on the individual studies and on the meta-analysis. The bias-adjusted meta-analysis estimates for RCT and MVR studies are similar: the pooled RR for RCTs is 1.04 (95% CI=0.89 to 1.23) and for MVR studies is 1.06 (95% CI=0.83 to 1.35), suggesting little evidence of an effect of beta-carotene on risk of CVD. The CIs for the individual studies and the meta-analysis are wider than for the bias unadjusted meta-analysis. The between-studies heterogeneity reduced after bias correction, likely due to this extra imprecision. The summary RR for all studies was 1.04 (95% CI=0.89 to 1.23) and no evidence for heterogeneity between study designs (p-value=0.87).

With a prior belief (**eTable 6**), similar results were obtained (**eFigure 3**); The summary RRs was 1.06 (95% CI=0.89 to 1.25) and no evidence for heterogeneity between study designs (p-value=0.97) (right panel of **eFigure 3**).

***eTable 7***: *Characteristics of included studies where endpoint is cardiovascular disease (CVD)*

| *First author* | *Study design* | *Outcome* | *Exposure measure* | *Median age (years)^* | *Follow-up (years)* | *Relative risk* (95% CI)* |
|---|---|---|---|---|---|---|
| Randomized controlled trials | | | | | | |
| Greenberg [8] | RCT (Skin Cancer Prevention Study) | CVD mortality | 50 mg/d | 63.2 | 8.2 (Dur. 4.3) | 1.13 (0.80, 1.58) |
| Green [9] | RCT (Nambour Skin Cancer Prevention Trial) | CVD mortality | 30 mg/d | 48.8 | 4.5 (Dur. 4.5) | 0.51 (0.19, 1.36) |
| Lee [10] | RCT (Women's Health Study) | All CVD events | 50 mg/2d | 54.6 | 4.1 (Dur. 2.1) | 1.14 (0.82, 1.59) |
| Hennekens [11] | RCT (Physicians' Health Study) | All CVD events | 50 mg/2d | 62.0 | 12.9 (Dur. 12) | 0.99 (0.91, 1.08) |
| | | CVD mortality | | | | 1.08 (0.93, 1.26) |
| Cook [12] | RCT (Women's Antioxidant Cardiovascular Study) | All CVD events | 50 mg/2d | 60.6 | 9.4 (Dur. 9.4) | 1.09 (0.96, 1.24) |
| | | CVD mortality | | | | 1.15 (0.95, 1.39) |
| *Conventional observational studies* | | | | | | |
| Genkinger [13] | Cohort Study (CLUE I and CLUE II) | CVD mortality | Dietary | 56.3 | 12.2 | 1.04 (0.68, 1.61) |

| | | | | | | |
|---|---|---|---|---|---|---|
| Stepaniak [14] | Cohort Study (Health, Alcohol and Psychosocial factors in Eastern Europe Study) | CVD mortality | Dietary | 58.3 | 7.23 | 0.96 (0.89, 1.02) |
| Buijsse [15] | Cohort Study (Zutphen Elderly Study) | CVD mortality | Dietary | 72.0 | 15 | 0.47 (0.24, 0.90) |
| de Oliveira Otto [16] | Cohort Study (Multi-Ethnic Study of Atherosclerosis) | All CVD events | Dietary | 61.8 | 6.2 | 0.92 (0.61, 1.38) |
| Fletcher [17] | Observational re-analysis of RCT (Medical Research Council Trial of Assessment and Management of Older People in the Community) | CVD mortality | Plasma | 78.5 | 4.4 | 0.79 (0.49, 1.28) |
| Bates [18] | Cohort study (National Diet and Nutrition Survey) | CVD mortality | Plasma | 76.6 | 13.5 | 0.92 (0.67, 1.31) |
| Goyal [19] | Cohort study (NHAMES III) | CVD mortality | Plasma | 44.8 | 14.2 | 0.96 (0.78, 1.17) |
| Greenberg [8] | Observational re-analysis of RCT (Skin Cancer Prevention Study) | CVD mortality | Plasma | 63.2 | 8.2 | 0.72 (0.55, 0.95) |
| Kilander [20] | Cohort study (Uppsala, Sweden) | CVD mortality | Plasma | 50.0 | 25.7 | 0.98 (0.70, 1.36) |
| Karppi [21] | Cohort study (Kuopio Ischaemic Heart Disease Risk Factor Study) | CVD mortality | Plasma | 57.8 | 15.9 | 0.66 (0.49, 0.89) |
| Mezzetti [22] | Cohort study (Val Vibrata Aging Project) | All CVD events | Plasma | 84.2 | 3.95 | 0.77 (0.42, 1.42) |
| Sesso [23] | Nested case-control study (embedded in Physicians' Health Study RCT) | All CVD events | Plasma | 69.7 | 2.1 | 0.93 (0.76, 1.15) |

CI, confidence interval ; Dur., intervention duration.
*Relative risk is extracted from systematic reviews, not individual studies.
^this varies between studies, some provide median, mean or range.

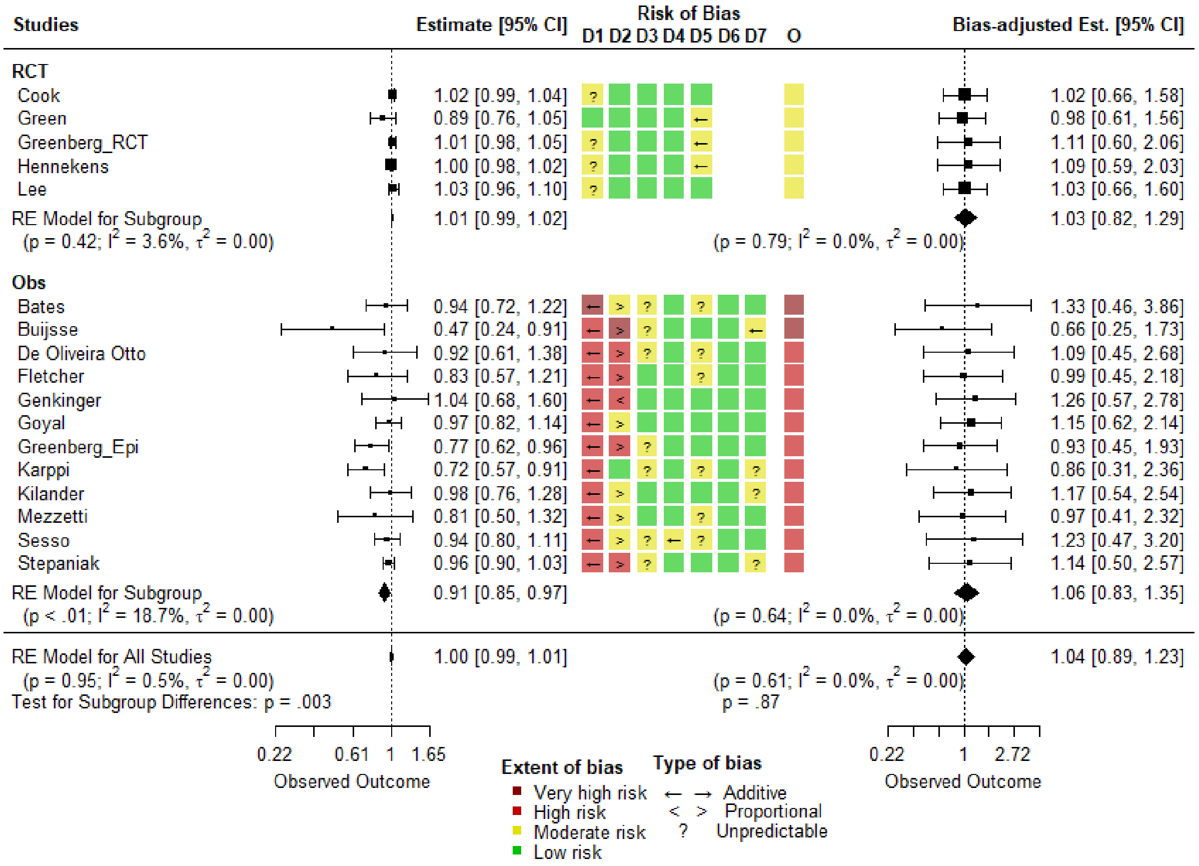

***eFigure 2**: Unadjusted (left panel) and bias-adjusted (right panel) random effects meta-analysis for CVD events as endpoint with risk-of-bias assessment (middle panel). See Table 1 for description of D1-D7 of each bias assessment tool. Priors is defined in Table 3. O is the overall judgment of risk of bias. RCT, randomized controlled trial; Obs, conventional observational study; CI, confidence intervals; p, p-value of the pooled effect estimate; $I^2$, heterogeneity variance / total variance; $\tau^2$, estimated amount of heterogeneity. The colours and symbols are shown in the legend on the plot.*

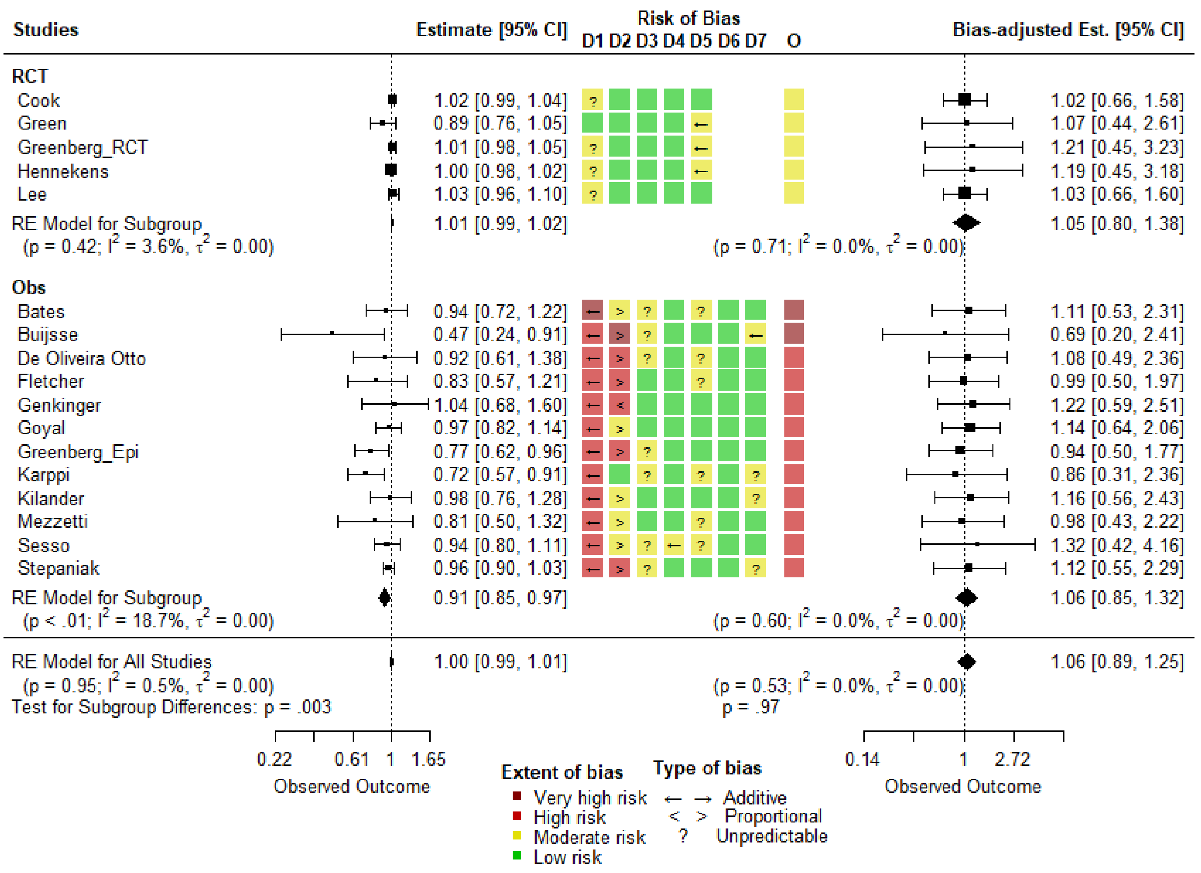

***eFigure 3:*** *Prior Sensitivity-Unadjusted (left panel) and bias-adjusted (right panel) random effects meta-analysis for CVD events as endpoint with risk-of-bias assessment (middle panel). See Table 1 for description of D1-D7 of each bias assessment tool. Priors is defined in* ***eTable 6****. O is the overall judgment of risk of bias. RCT, randomized controlled trial; Obs, conventional observational study; CI, confidence intervals; p, p-value of the pooled effect estimate;* $I^2$*, heterogeneity variance / total variance;* $\tau^2$*, estimated amount of heterogeneity. The colours and symbols are shown in the legend on the plot.*
